# Semi-automatic segmentation of the fetal brain from Magnetic Resonance Imaging

**DOI:** 10.1101/2022.03.10.22272216

**Authors:** Jianan Wang, Emily S. Nichols, Megan E. Mueller, Barbra de Vrijer, Roy Eagleson, Charles A. McKenzie, Sandrine de Ribaupierre, Emma G. Duerden

**Author notes:** **Address correspondence to:** Emma G. Duerden, Faculty of Education, 1137 Western Rd, London, Ontario N6G 1G7. These authors contributed equally (Sandrine de Ribaupierre, Emma G. Duerden).

## Abstract

Template-based segmentation techniques have been used for targeting deep brain structures in fetal MR images. In this study, two registration algorithms were compared to determine the optimal strategy of segmenting subcortical structures in T2-weighted images acquired during the third trimester of pregnancy. Adult women with singleton pregnancies (n=9) ranging from 35-39 weeks gestational age were recruited. Fetal MRI were performed on 1.5 T and 3 T scanners. Automatic fetal brain segmentation and volumetric reconstruction algorithms were performed on all subjects using the NiftyMIC software. An atlas of cortical and subcortical structures (36 weeks’ gestation) was registered into native space using ANTs (Automatic Normalization Tools) and FLIRT (FMRIB’s linear image registration tool). The cerebellum and thalamus were manually segmented. Dice coefficients were calculated to validate the reliability of automatic methods and to compare the performance between ANTs (nonlinear) and FLIRT (affine) registration algorithms compared to the gold-standard manual segmentations. The Dice-kappa values for the automated labels were compared using the Wilcoxon test. Comparing cerebellum and thalamus masks against the manually segmented masks, the median Dice-kappa coefficients for ANTs and FLIRT were 0.76 (interquartile range [IQR]= 0.56-0.83) and 0.65 (IQR=0.5-0.73), respectively. The Wilcoxon test (Z=4.9, *P*<0.01) indicated that the ANTs registration method performed better than FLIRT for the fetal cerebellum and thalamus. We found that a nonlinear registration method, provided improved results compared to an affine transformation. Nonlinear registration methods may be preferable for subcortical segmentations in MR images acquired in third-trimester fetuses.

## Introduction

The fetal brain undergoes exponential development in the third trimester of pregnancy. With advances in fetal magnetic resonance imaging (MRI), research in neonatal neuroscience has shifted to identify in utero predictors of fetal brain dysmaturation. Better understanding of the trajectories of fetal brain development through volumetric segmentation can aid in functional outcome prediction in at-risk neonates.

Template-based segmentation techniques have been used to target deep brain structures in fetal MRI images. Landmark-based rigid image transformation has been applied for volumetric and cortical measures [1]. However, landmark-based registration can be time consuming and require manual editing.

In this study, a platform of novel fetal brain segmentation and volumetric reconstruction algorithms were applied to T2-weighted fetal MR images acquired during the third trimester [2]. Two different automatic registration tools, affine and nonlinear atlas registration algorithms, were applied to the reconstructed images and compared to determine an optimal fetal subcortical segmentation strategy. Volumetric segmentations of the fetal subcortical structures can be used to identify brain-based biomarkers for perinatal health [1]. Identifying early predictors of brain dysmaturation is critical for developing antenatal treatment strategies to better support fetal neurodevelopment and neonatal outcomes.

## Methods

### Participants and MRI protocol

Eight pregnant adult women ranging from 35 to 39 weeks of gestational age participated. The participants were scanned on either a 1.5T or 3T MRI scanner. T2-weighted anatomical single shot fast spin echo (SSFSE) images (repetition time [TR] > 1200 msec, echo time [TE]: 81.36-93.60 msec, voxel size 0.98*1.96*8 mm3 and 0.125*0.17*9 mm3) [3] were acquired for fetuses.

### MR image preprocessing

Motion artifacts in fetal MR images are inevitable. The NiftyMIC platform with fetal brain segmentation [4] and volumetric reconstruction algorithms [2] was utilized to mitigate the motion artifacts’ effects on deep brain structure estimation. The fetal brain segmentation algorithm of fetal_brain_seg application from NIftyMIC using a 2D convolutional neural-network (CNN), specifically the 2D P-Net [2,4] was performed on all subjects. The outcome segmented low-resolution 2D slices of fetal brain masks in coronal, axial, and sagittal orientations were taken as input for the NiftyMIC volumetric reconstruction process. Orientation tags were manually adjusted according to the fetal brain atlas. Next, thalamus and cerebellum segmentations on the reconstructed and reoriented fetal brain volumes were manually performed by one of the authors (MM). The manually labeled masks were considered as ground truth for later atlas registration algorithms comparison.

### Atlas registration

Nonlinear atlas registration was performed using ANTs [5] that came with the well-described SyN (symmetric image normalization) method [6] and linear (affine) atlas registration was performed using FLIRT [7,8]. The fetal brain atlas (Fig. 1) is an averaged 36-weeks GA template with pre-defined labels of vital deep brain structures including the thalamus and cerebellum. The atlas was nonlinearly and linearly registered into the native participant 3D MRI space. The transformed atlas labels were used as thalamus and cerebellum masks and were compared with manual masks by calculating Dice Kappa coefficients for the accuracy test.

**Fig. 1.**
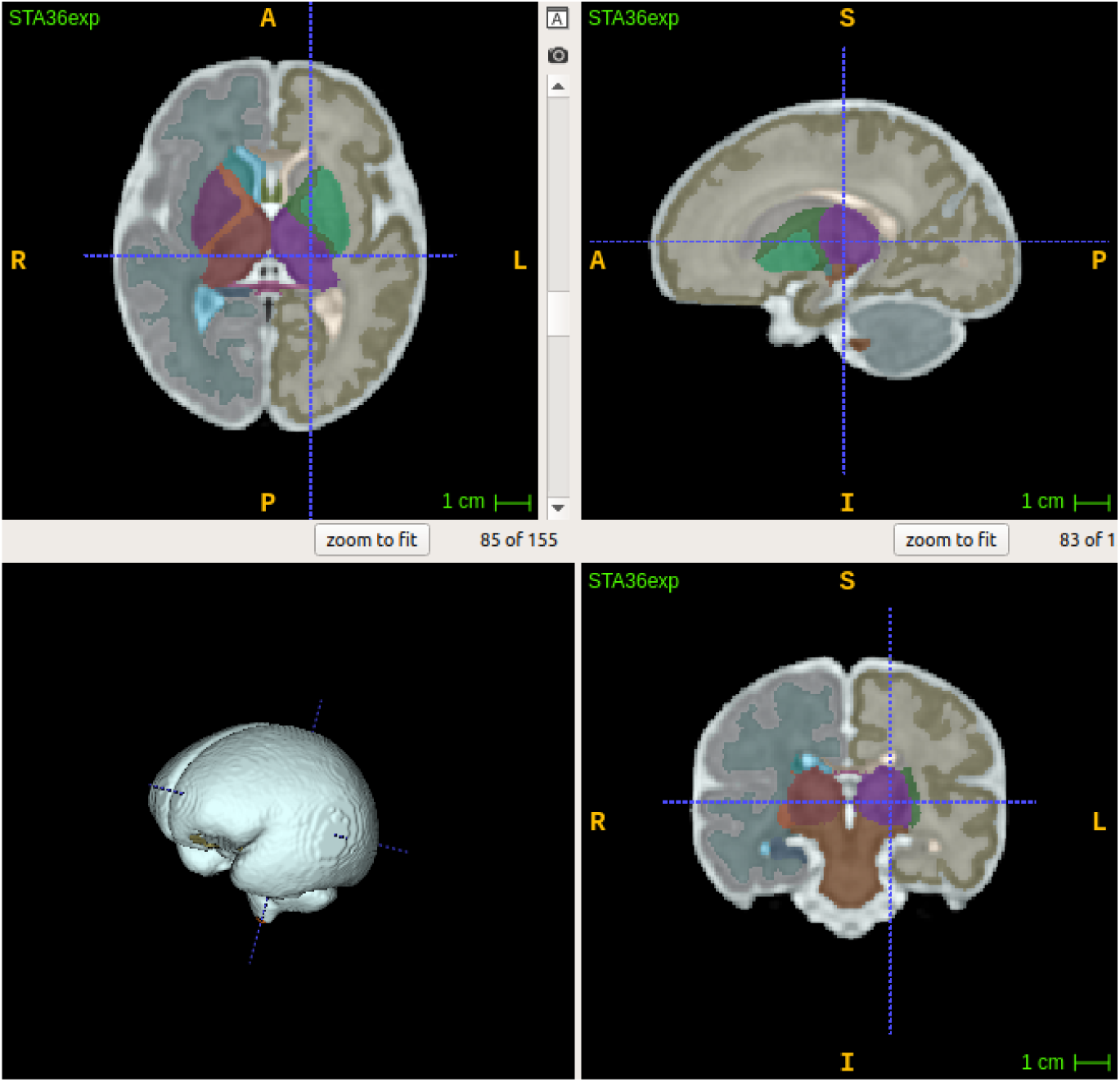
Fetal regional tissue-based atlas (36 weeks’ GA)

### Statistical Analysis

Statistical analyses were performed using SPSS (v 26, Armonk, NY). The resulting Dice Kappa values were non-normally distributed. Therefore, a nonparametric Wilcoxon signed ranks test for paired data was applied to the Dice Kappa Values. An alpha level of p<0.05 was selected.

## Results

The 2D fetal brain masks of the 2D stacks of the original fetal brain MR images were segmented by fetal_brain_seg in axial, coronal, and sagittal planes. Then the fetal brain masks, which are shown in Fig. 1, were manually adjusted for over- and under-estimations by automatic segmentation algorithm from fetal_brain_seg for optimal performance of NiftyMIC’s volumetric reconstruction algorithm. The 2D masks were then reconstructed into 3D volumes (Fig. 2) by NiftyMIC. Based on the reconstructed volume, the cerebellum and thalamus (Fig. 3) volumes were manually segmented. The template-based automatic cerebellum and thalamus segmentations were performed by FLIRT and ANTs (Fig. 3).

**Fig. 2.**
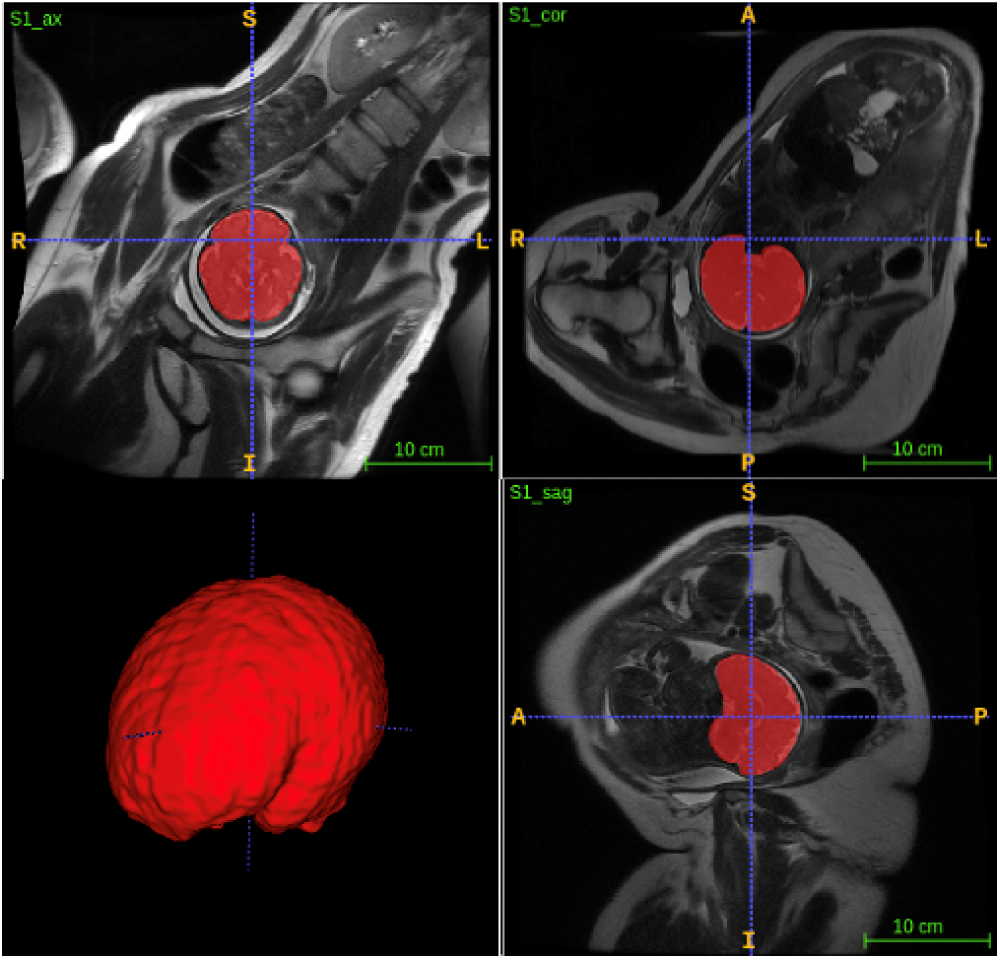
The stacks of 2D T2 weighted fetal MR image for a representative participant. Red labels are the fetal brain masks in axial, coronal, and sagittal planes from left to right. The volumetric reconstruction result of a representative subject using the NiftyMIC algorithm.

**Fig. 3.**
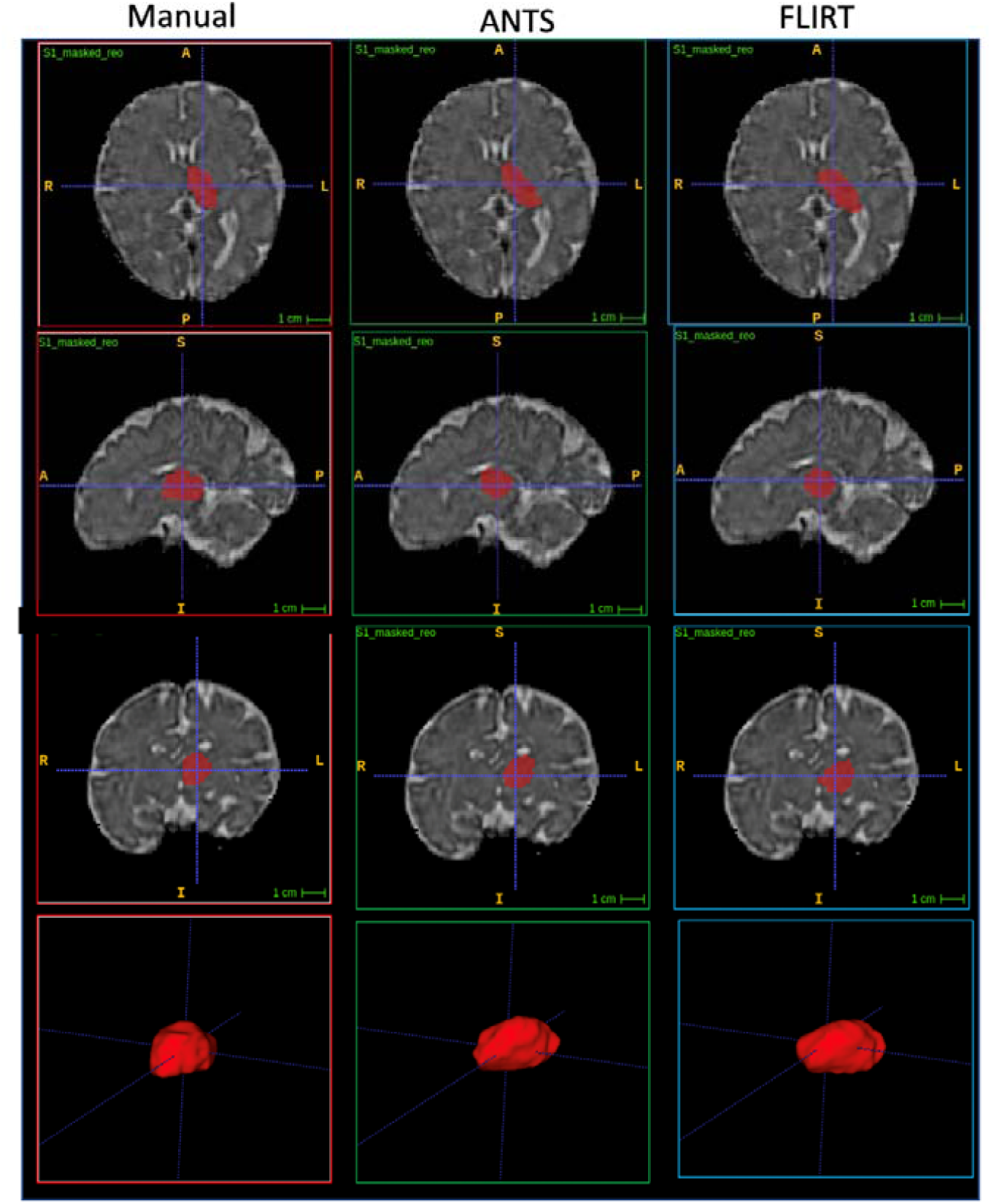
Segmented left thalamus on a reconstructed MRI of one representative participant using manual segmentation (left), ANTs (middle), and FLIRT (right).

The comparison of FLIRT and ANTs was performed by comparing the commonly overlayed areas of both image registration tools’ masks against the manual masks using the convert3D toolkit from ITK_SNAP. Dice Kappa coefficients representing the comparisons of the bilateral cerebellar hemispheres are shown in Table 1, and that of the thalamus (bilaterally) are shown in Table 2. The median of all Dice Kappa values for the ANTs-registered cerebellum and thalamus masks aligning against the manual masks was 0.77 (interquartile range [IQR]=0.56-0.83) and that for the FLIRT-registered masks is 0.66 (IQR=0.5-0.73) (Table 3). Wilcoxon signed ranks test was computed to compare the statistic differences between the repeated-measures Dice Kappa values and the ANTs-registered images had significantly higher coefficients (z= 4.9, p ≤ 0.01).

**Table 1.**
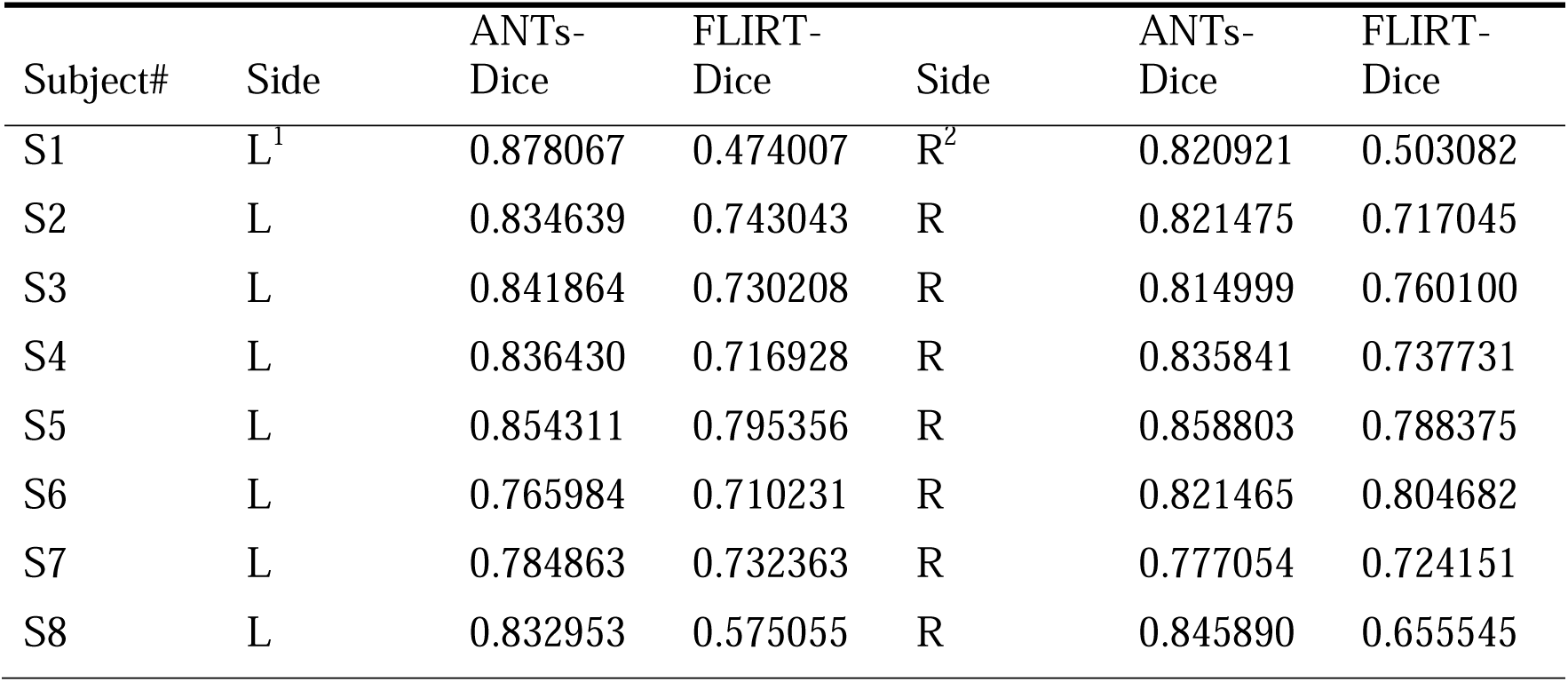
Dice Kappa coefficients for left and right cerebellum (ANTs vs. FLIRT).

**Table 2.** Dice Kappa coefficients for left and right thalamus (ANTs vs. FLIRT)

| Subject# | Side | ANTs-Dice | FLIRT-Dice | Side | ANTs-Dice | FLIRT-Dice |
| --- | --- | --- | --- | --- | --- | --- |
| S1 | L | 0.663834 | 0.613340 | R | 0.816123 | 0.691856 |
| S2 | L | 0.383442 | 0.359765 | R | 0.465639 | 0.441790 |
| S3 | L | 0.214058 | 0.206838 | R | 0.225466 | 0.188555 |
| S4 | L | 0.708085 | 0.689478 | R | 0.478793 | 0.429654 |
| S5 | L | 0.631239 | 0.620896 | R | 0.750000 | 0.699609 |
| S6 | L | 0.519163 | 0.486708 | R | 0.550265 | 0.528596 |
| S7 | L | 0.570672 | 0.549439 | R | 0.413926 | 0.385913 |
| S8 | L | 0.691663 | 0.658274 | R | 0.649737 | 0.558755 |

**Table 3.** Median, mean, and standard deviation of all Dice values

| Heading level | Median | Standard deviation |
| --- | --- | --- |
| ANTs vs. manual | 0.772 | 0.686 |
| FLIRT vs. manual | 0.657 | 0.602 |

## Conclusion

Fetal brain MR images were successfully segmented and reconstructed into 3D subcortical volumes. The affine registration algorithm (FLIRT) and nonlinear registration algorithm (ANTs) were successfully applied and compared to manually segmented masks. The median Dice Kappa value of the ANTs-registered masks (vs. manual masks) was higher than that of the FLIRT-registered masks (vs. manual masks). Thus, the ANTs nonlinear registration method produced improved template-based segmentations of fetal subcortical brain structures.

The nonlinear registration method performed better on our dataset for two main reasons. Firstly, the number of degrees of freedom of the deformation is correlated to registration accuracy. SyN that is used in ANTs has millions of degrees of freedom [9], which is much more than that of the affine registration offered by FLIRT (up to 12 degrees of freedom for 3D-to-3D model). Secondly, is that the reconstructed fetal MR images still include intrasubject motion artifacts. In turn, the nonlinear registration method may be helpful when data include nonlinear image distortions.

As this study has included limited number of participants, the study of brain-based biomarkers that associate growth restriction with higher risk of altered neurodevelopment outcome [10,11] is still ongoing. This presented workflow of different toolkits using deep learning and nonlinear image transformation algorithms can help to better characterize third-trimester fetal brain development to achieve this goal.

## Data Availability

All data produced in the present study are available upon reasonable request to the authors.

## Acknowledgments

The authors would like to thank David Reese and Dr. Trevor Wade for MRI technical support. The Canadian Institutes of Health Research, New Frontiers in Research Fund and the Canada First Research Excellence Fund provided the funding for this research. We also sincerely thank the women and their families who participated in this study.

## Footnotes

1 Left

2 Right

## References

1. Wu Y, Lu Y-C, Jacobs M, Pradhan S, Kapse K, Zhao L, et al. Association of Prenatal Maternal Psychological Distress With Fetal Brain Growth, Metabolism, and Cortical Maturation. JAMA Netw open. 2020;3: e1919940–e1919940. doi:10.1001/jamanetworkopen.2019.19940

2. Ebner M, Wang G, Li W, Aertsen M, Patel PA, Aughwane R, et al. An automated framework for localization, segmentation and super-resolution reconstruction of fetal brain MRI. Neuroimage. 2020;206. doi:10.1016/j.neuroimage.2019.116324

3. Goldberg E, McKenzie CA, de Vrijer B, Eagleson R, de Ribaupierre S. Fetal Response to a Maternal Internal Auditory Stimulus. J Magn Reson Imaging. 2020; 1–7. doi:10.1002/jmri.27033

4. Ebner M, Wang G, Li W, Aertsen M, Patel PA, Aughwane R, et al. An Automated Localization, Segmentation and Reconstruction Framework for Fetal Brain MRI. Medical Image Computing and Computer Assisted Intervention. Cham; 2018. pp. 313– 320.

5. Avants B, Tustison N, Johnson H. Advanced Normalization Tools (ANTS). Insight J. 2009; 1–35.

6. Avants BB, Epstein CL, Grossman M, Gee JC. Symmetric diffeomorphic image registration with cross-correlation: Evaluating automated labeling of elderly and neurodegenerative brain. Med Image Anal. 2008;12: 26–41. doi:https://doi.org/10.1016/j.media.2007.06.004

7. Jenkinson M, Smith S. A global optimisation method for robust affine registration of brain images. Med Image Anal. 2001;5: 143–156. doi:https://doi.org/10.1016/S1361-8415(01)00036-6

8. Jenkinson M, Bannister P, Brady M, Smith S. Improved Optimization for the Robust and Accurate Linear Registration and Motion Correction of Brain Images. Neuroimage. 2002;17: 825–841. doi:https://doi.org/10.1006/nimg.2002.1132

9. Klein A, Andersson J, Ardekani BA, Ashburner J, Avants B, Chiang M-C, et al. Evaluation of 14 nonlinear deformation algorithms applied to human brain MRI registration. Neuroimage. 2009;46: 786–802. doi:10.1016/j.neuroimage.2008.12.037

10. Nosarti C, Murray RM, Hack M, editors. Neurodevelopmental Outcomes of Preterm Birth. Cambridge: Cambridge University Press; 2010. doi:10.1017/CBO9780511712166

11. Blencowe H, Lee ACC, Cousens S, Bahalim A, Narwal R, Zhong N, et al. Preterm birth-associated neurodevelopmental impairment estimates at regional and global levels for 2010. Pediatr Res. 2013;74: 17–34. doi:10.1038/pr.2013.204

